# Tumour volume distribution can yield information on tumour growth and tumour control

**DOI:** 10.1101/2020.09.01.20185595

**Authors:** Uwe Schneider, Jürgen Besserer

## Abstract

**Background:** It is shown that tumour volume distributions, can yield information on two aspects of cancer research: tumour induction and tumour control.

**Materials and methods:** From the hypothesis that the intrinsic distribution of breast cancer volumes follows an exponential distribution, firstly the probability density function of tumour growth time was deduced via a mathematical transformation of the probability density functions of tumour volumes. In a second step, the distribution of tumour volumes was used to model the variation of the clonogenic cell number between patients in order to determine tumour control probabilities for radiotherapy patients.

**Results:** Distribution of lag times, i.e. the time from the appearance of the first fully malignant cell until a clinicaly observable cancer, can be used to deduce the probability of tumour induction as a function of patient age. The integration of the volume variation with a Poisson-TCP model results in a logistic function which explains population-averaged survival data of radiotherapy patients.

**Conclusions:** The inclusion of tumour volume distributions into the TCP formalism enables a direct link to be deduced between a cohort TCP model (logistic) and a TCP model for individual patients (Poisson). The TCP model can be applied to non-uniform tumour dose distributions.

## Introduction

The estimation and analysis of tumour size^†^ and associated size distributions have mainly been applied to modelling their effects on cancer survival or tumour progression [1–8]. A major conclusion of published work is that tumour size has a significant impact on survival of patients treated with radiotherapy. In this work we focus on alternative applications of tumour size distributions. We will show that tumour-size distributions, or more precise tumour-volume distributions, can yield valuable information on two completely different areas of cancer research. Firstly we will show that the probability density distribution of detected tumour sizes is strictly related to the probability density function of tumour growth time. The tumour growth time, often called lag time, is the time from the appearance of the first fully malignant cell until development of a clinical observable cancer. In tumour induction models this lag time is usually estimated as a fixed number. We believe that the use of the determined lag time distribution in tumour induction models has the potential to improve the quality of such models.

Secondly, the distribution of cancer volumes reflects the distribution of clonogenic cell numbers (assuming a constant clonogenic cell density) and thus can be used for modelling tumour control probability for cohorts of radiotherapy cancer patients. Currently no general analytical formulation of TCP exists which includes the population variation of tumor volumes. Often a logistic function is used to model population averaged TCP [9], however, to our knowledge no strict derivation of a logistic TCP model exists. We will show that the probability density function of cancer sizes together with a Poisson based TCP model yields an analytic expression for TCP in terms of a logistic function.

## Materials and Methods

### Probability distributions of tumour volumes

We assume here that the intrinsic probability density function (pdf) for the distribution of tumour volumes follows an exponential distribution.

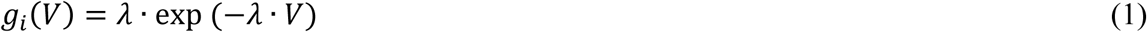

As the mean value of an exponential distribution is 1/*λ* one can write:

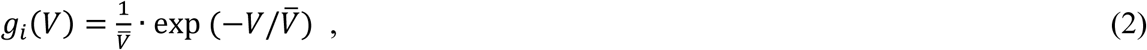

where 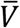is the average tumour volume.

With intrinsic we mean here that the exponential probability distribution describes the distribution of all tumour volumes in a cohort of subjects, independently of a potential diagnosis. E.g., if we would select a specific age group of women, all breast tumours, including also unobserved and microscopically small ones, would be distributed exponentially. However, the mean tumour volume is assumed to change with the age of the observed cohort.

Obviously, such an intrinsic probability density function can never be observed, as tumours are only detected after they reach a certain volume. For tumour diagnosis a certain minimal tumour size is required such that they are detectable with state-of-the–art diagnostic procedures. If it is assumed that for small volumes the detection probability of a tumour is increasing linearly with volume, the actually observed tumour volume distribution *g*_*0*_*(V)* can be written as:

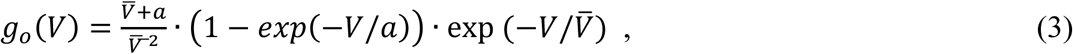

where the term 1 − *exp*(−*V/a*) is for small volumes *V/a*. The normalization constant was chosen such that the integral over the distribution function *g*_*0*_ is 1. In equation 3, *a* is a parameter representing a characteristic volume such that (1 − *exp*(−*V/a*) describes the detection rate. In the limit of *a = 0* all tumours are detected and the distribution function describes the intrinsic tumour-volume distribution. The value of *a* depends on the quality of the applied diagnostic procedures. As we will see later, *a* was decreased after mammography was employed for the detection of breast cancers.

For model testing, breast cancer volume distributions were selected, as they include usually a large number of patients and the data are available at different times, which allows the impact of improved diagnosis on the parameter *a* to be studied. Here we use the data of Carter et al. [10] and Rosenberg et al. [11]. The study of Carter et al. [10] included 24,740 cases recorded in the Surveillance, Epidemiology, and End Results (SEER) Program of the National Cancer Institute between 1977 and 1982. Rosenberg et al. [11] collected data from 72,367 cases from 1973 to 1998 also using the SEER data base. Equation 3 was fitted to the epidemiological data by using the least squares method. The fit was performed such that the average volume 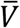was kept the same for both data-sets, assuming that the intrinsic volume distribution is identical for both data-sets. The parameter *a* was fitted separately, as the observation interval for the two data-sets was different.

### Determination of the probability distribution of tumour growth times

Many tumour induction models [12–16] assume that pre-malignant stem cells transform themselves with a certain probability, into fully malignant cells, and, after a certain lag time *L*, into a clinically observable cancer. Thus, *L* is the lag (time) between the first fully malignant cell and cancer. Usually the parameters of tumour induction models are determined by fitting tumour incidence data by assuming a fixed lag time usually estimated from the A-bomb survivor data [17–18] or from comparing tumour size at different time [19].

If a simple exponential tumour growth model is used to describe the size increase of the tumour during the lag time, the pdf which describes the tumour volume distribution *g*_*i*_*(V)* can be transformed into the pdf of the lag time *f(L)*. Thus, knowledge of the tumour volume distribution function can yield the shape of the tumour lag time distribution and vice versa. A schematic concept of the process is provided in Figure 1.

**Figure 1.**
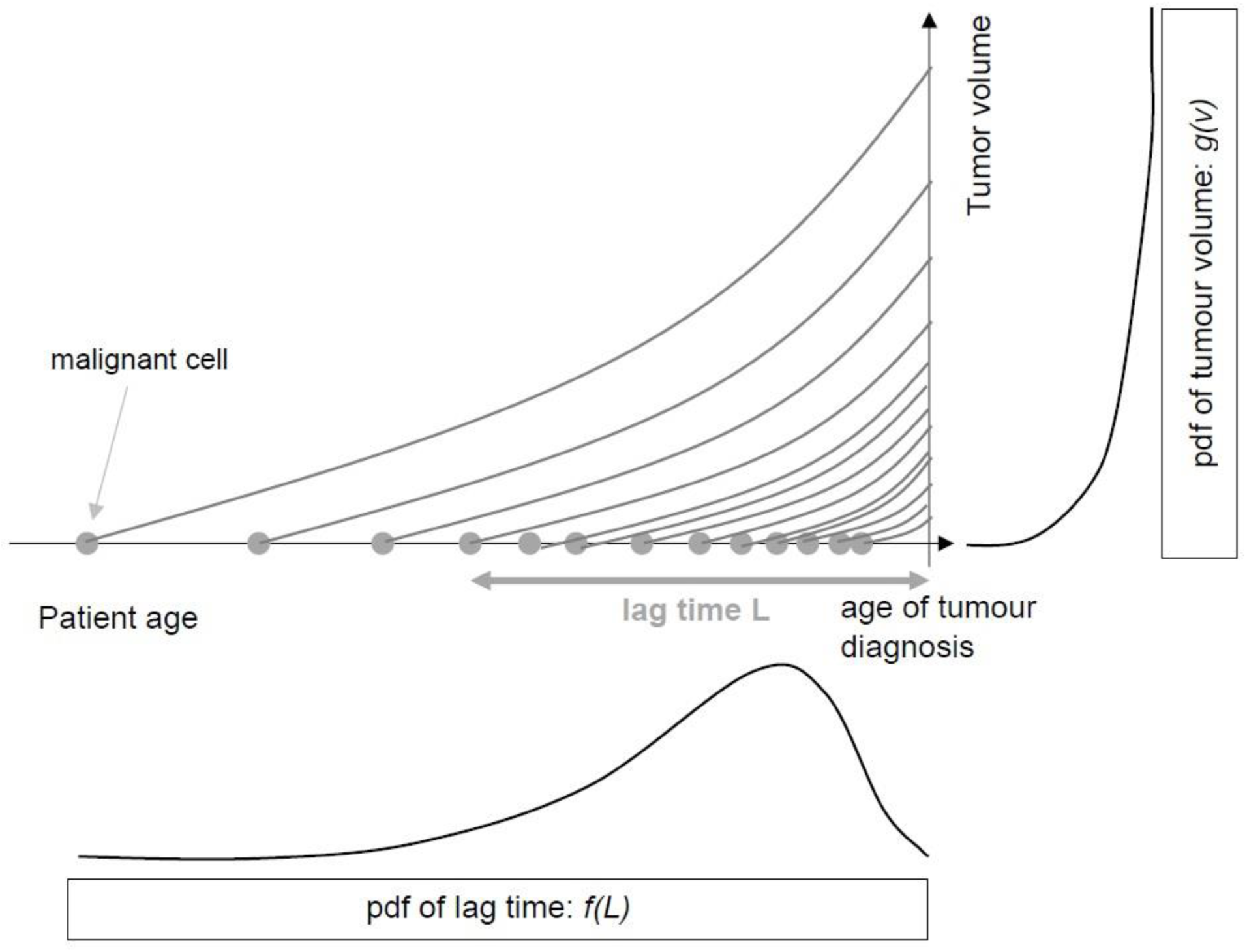
A schematic concept of the relation between the tumour volume pdf and the tumour growth time pdf. The horizontal axis shows tumour induction as a function of age. The occurrence of malignant cells is displayed with gray circles. Each cell grows exponentially (shown by the lines) and after a certain lag time a clinical observable tumour is grown. The clinical observable tumours are distributed according to plot shown on the vertical axis (equation 2), the corresponding lag times distributions are shown on the horizontal axis.

Here an exponential tumour growth function is used converts one probability density function into another:

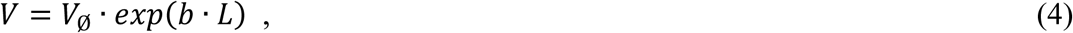

where *b* is the tumour growth factor and *L* the lag time. The volume *V*_*Ø*_ is the initial volume before exponential growth starts. Here *V*_*Ø*_ = 10^−9^ cm^3^, i.e., the volume of one cell, was used. The transformation between the two pdfs is a strict mathematical procedure given by [20]:

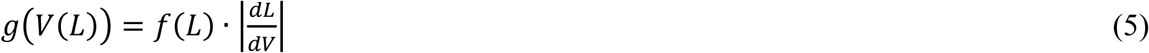

Using equation (4) we get:

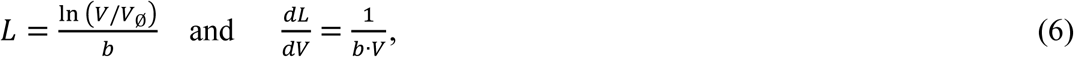

with *g(V)* from equation (2), *f(L)* can be easily determined using equations 5 and 6:

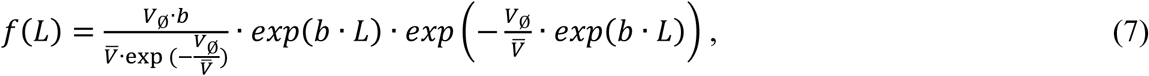

where equation 7 was normalized such that the integral over *f(L)* from *L =* 0 to infinity is unity. We have chosen infinity as the upper integration limit instead of a realistic maximum tumour volume, as the probability for very large tumours is practically zero. E.g., assuming an average tumor volume of 40 cm^3^ an integration of the exponential distribution up to a realistic upper limit of 500 cm^3^ yields 0.999996 instead of 1.

For the visualization of a realistic lag time distribution we have used the tumour growth factor *b =* 1.05 y^−1^ for breast cancer taken from Talkington and Durrett [19].

### Determination of TCP

Another interesting application of the exponential pdf of tumour volumes described by equation 2 is the application to models of tumour control probability (TCP) following radiotherapy. Most TCP models are based on a calculation of the Poisson probability of there being no surviving cells in a population of tumours after a fractionated treatment [21, 22]:

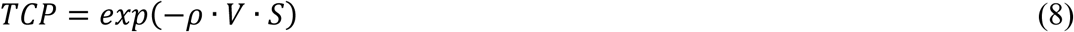

where *ρ* is the number of tumour clonogens per unit volume, *V* is the tumour volume and *S* is the surviving fraction of cells after radiotherapy treatment. If the TCP curves are generated with single values of intrinsic radiosensitivity and clonogenic cell number, the experimentally obtained data cannot be fitted, as the TCP curves show a very steep gradient. This discrepancy is usually explained by the fact that TCP curves derived from populations of human tumours that have been similarly treated include different tumour volumes, different initial clonogen densities, different intrinsic radiosensitivities and different oxygenation status. However, these parameters cannot all be modeled analytically and as a consequence, they are often varied numerically (e.g. [23, 24]). An alternative to numerical calculations is the use of a logistic model to fit the observed tumour control (e.g. [9]). However, to our knowledge it has never been shown that this empirical approach is based on a mechanistic understanding of the involved processes.

By assuming that the tumour volumes are distributed according to equation 2, and assuming a constant clonogenic density we obtain:

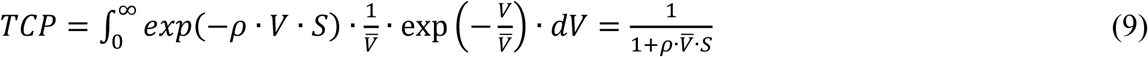

The result of this integration yields an analytic expression for TCP in terms of a logistic model were survival *S* is modelled by an exponential function using the linear quadratic model:

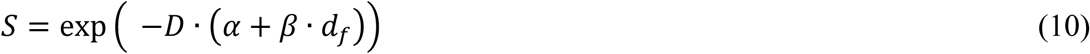

Some typical parameters have been used: *ρ* = 10^8^ cm^−3^, 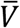 = 10, 100 and 1000 cm^3^, *α* = 0.36 Gy^−1^, *β* = 0.036 Gy^−2^, *D* = 30…70 Gy and *d*_*f*_ = 2 Gy, where *α* and *β* are the parameters of the LQ-model, *D* is the total radiotherapy dose and *d*_*f*_ is the dose per fraction.

## Results

### Probability distributions of tumour sizes

In Figure 2 tumour volume distributions of breast cancer are shown. The data points marked by diamonds represent the volume distribution from the Carter data including cases until 1982, whereas the squares belong to the Rosenberg data obtained between 1973 and 1998. The statistical error is of the order of the size of the symbols. The data were fitted to equation 3. A shift of the maximum of the probability distribution of the newer data to smaller volumes can clearly be observed. This is reflected by a smaller value of *a* = 0.42 cm^3^, hence better detection efficiency, when compared to older data (*a* = 1.84 cm^3^). The average volume obtained from the fit of the exponential distribution to the two data sets is 20.7 cm^3^. The fitted parameters with the corresponding errors are listed in table 1.

**Table 1.**
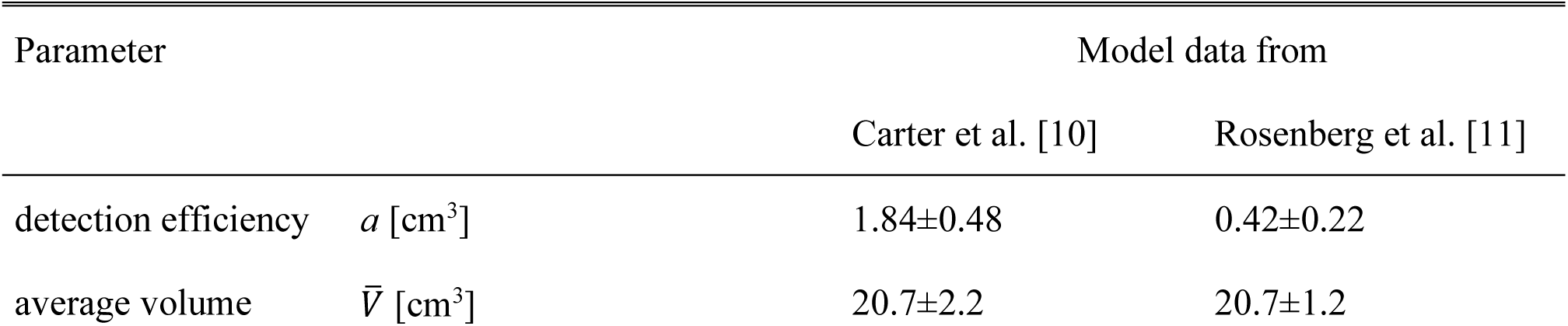
Fitted parameters of the for tumour volume distributions with one standard deviation.

| Parameter |  | Model data from |  |
| --- | --- | --- | --- |
|  |  | Carter et al. [10] | Rosenberg et al. [11] |
| detection efficiency | $a$ [cm<sup>3</sup>] | 1.84±0.48 | 0.42±0.22 |
| average volume | $\bar{V}$ [cm<sup>3</sup>] | 20.7±2.2 | 20.7±1.2 |

**Figure 2.**
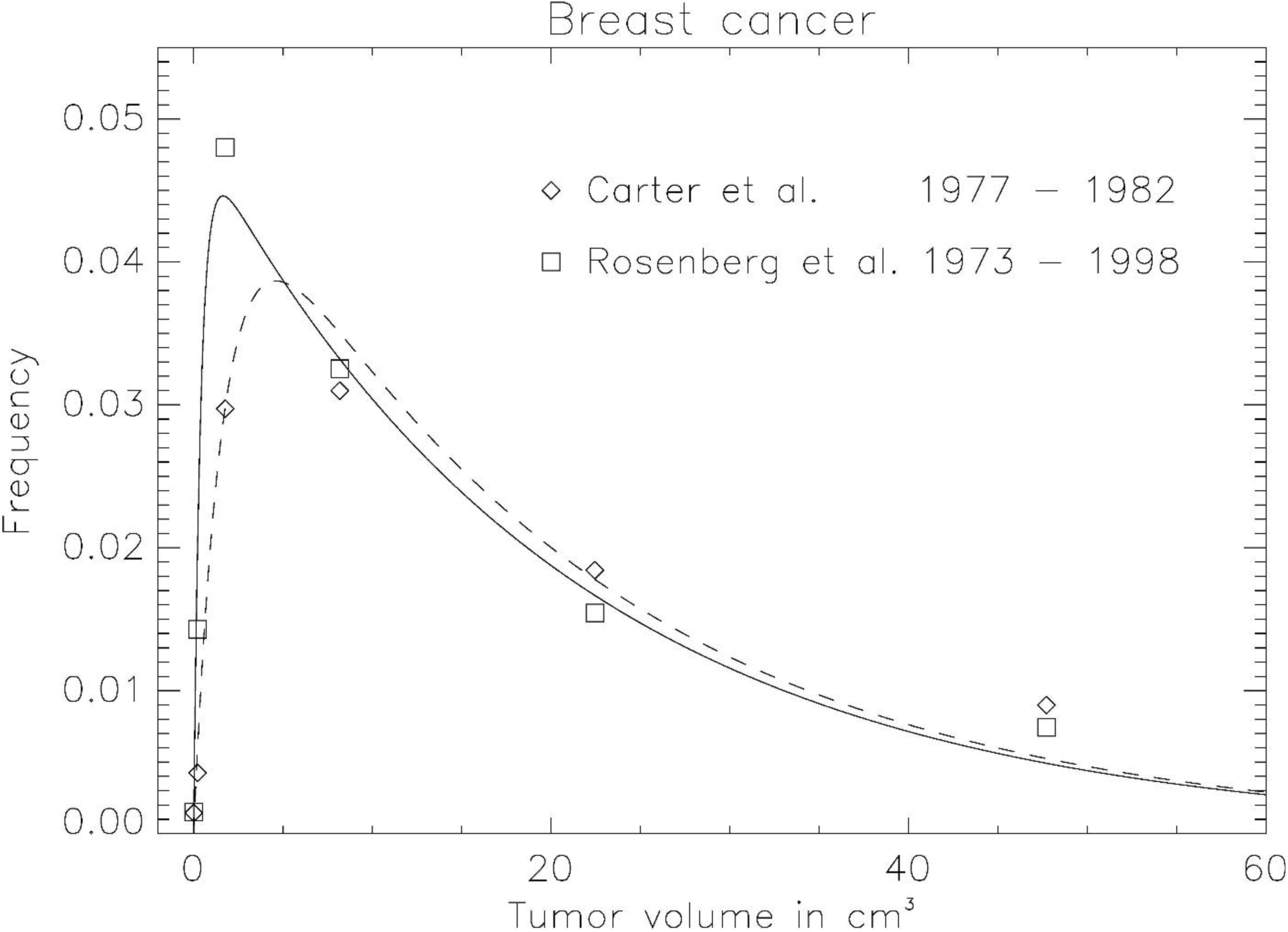
Probability density distribution of breast cancer induction. The diamonds and squares show the frequency distribution of the observed data from Carter et al. [10] and Rosenberg et al. [11], respectively. The lines represent the fit of equation 3. The fitted parameters *a* and 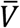 are listed in table 1.

### Determination of the probability distribution for tumour growth times

The expression for the pdf of breast tumour lag times from equation 7 is plotted in Figure 3. A typical tumour growth factor *b* for breast cancer and an average tumour volume of 20.7 cm^3^ obtained from the comparison of the tumour size distribution with the epidemiological data was used. The average lag time for breast cancer was determined to be 22.1 years with a variance of 1.5 years. This value is consistent with the findings from the A-bomb survivors with a lag time between 10 and 30 years [17–18] and with studies comparing tumour size at different time. Talkington and Durrett [20] found for breast cancer a growth rate of 1.05/year for an exponential model. This translates into a lag time of 22 years for a tumor of volume 1.3 cm^3^.

**Figure 3.**
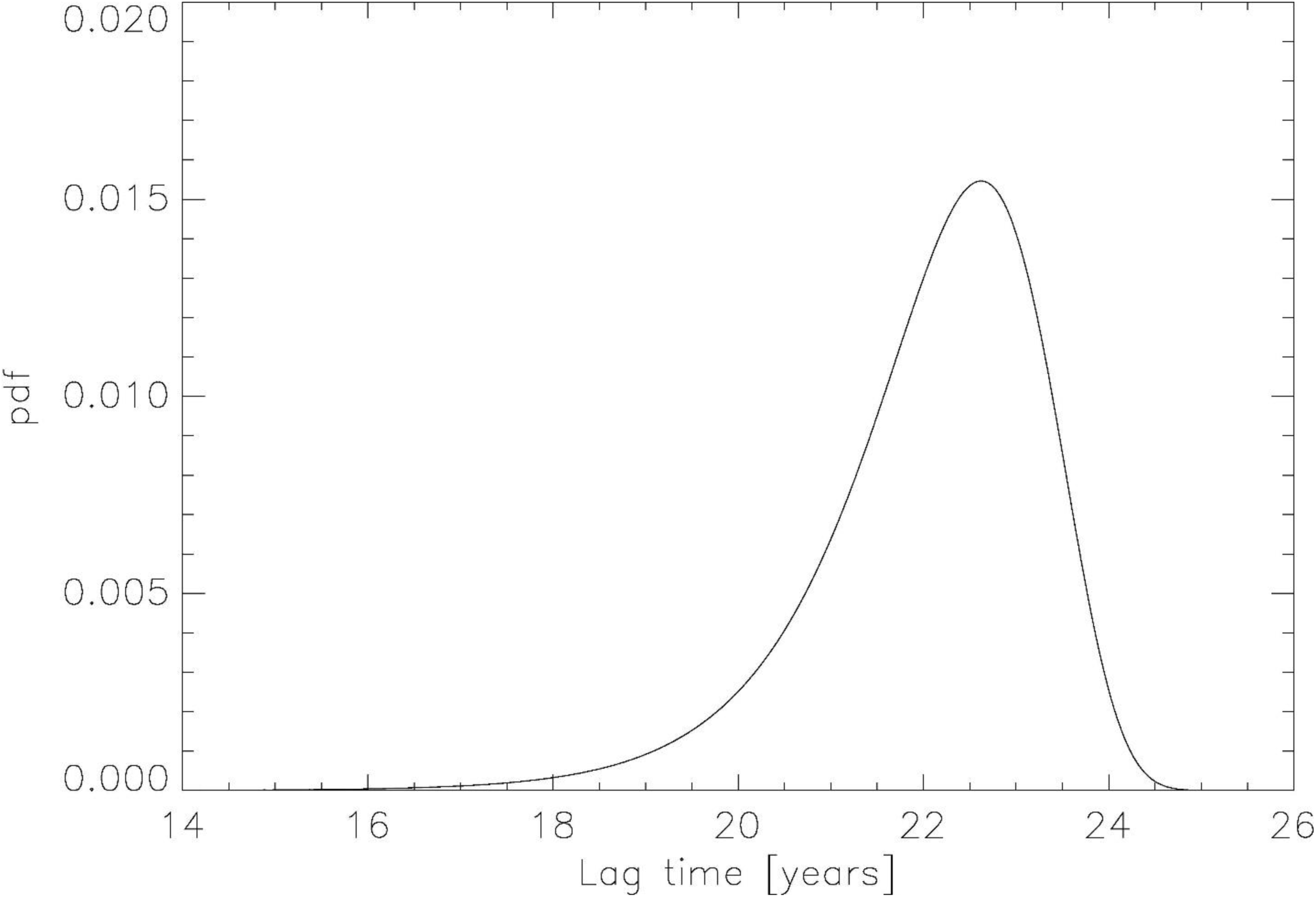
Plot of the pdf of breast tumour lag times from equation 7. A typical tumour growth factor *b = 1*.*05* for breast cancer and an average tumour volume of 20.7 cm^3^ was used.

### Determination of TCP

In Fig. 4 the TCP model of equation 8 is plotted as the dotted line for three different average tumour volumes 10, 100 and 1000 cm^3^, respectively. The solid lines represent the TCP calculations including an exponential distribution of tumour volumes from equation 9. Clearly, a broadening of the TCP curve is visible.

**Figure 4.**
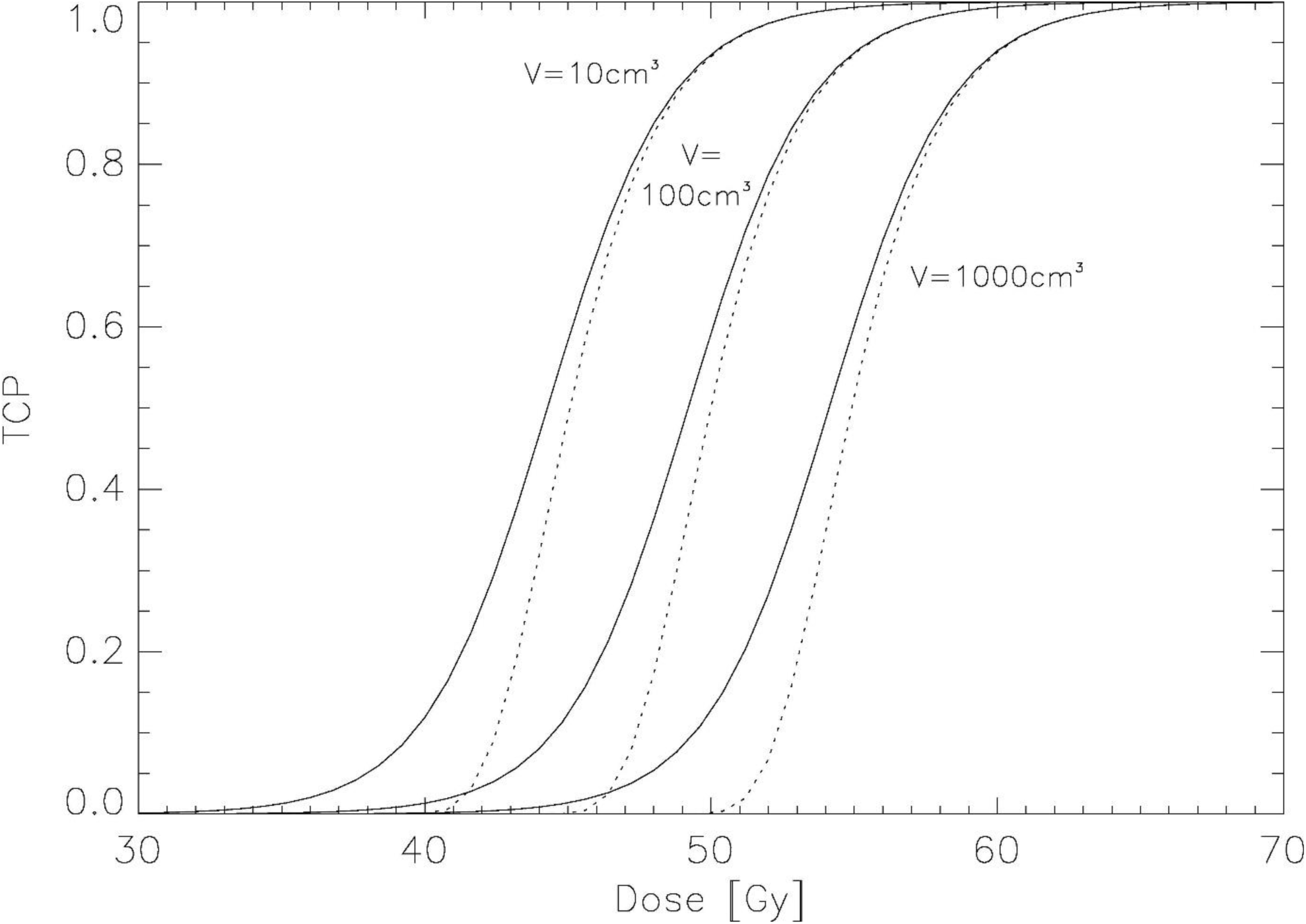
Plot of the TCP model of equation 8 for individual patients as the dotted line for three different average tumour volumes 10, 100 and 1000 cm3, respectively. The solid lines shows the TCP model for a volume averaged patient cohort according to equation 9.

## Discussion

In this work it was hypothesized that the intrinsic distribution of tumour volumes in a cohort is exponentially distributed. To account for the fact that the probability of the clinical detection of a tumour increases with tumour volume, the exponential distribution was modified with a sigmoidal function. The fit of this distribution function to breast volume data from literature covering two different time intervals of breast cancer diagnosis results in two different locations of the distribution maximum and consequently in different values of parameter *a*. The data set covering diagnosed breast cancers from 1973 – 1998 results in a much smaller *a* (0.31) than the older data set (1977 - 1982; *a=*1.20). This can be explained by the clinical introduction of mammography in the late 1980s which resulted in the diagnosis of much smaller breast tumours than before [8]. Therefore, a smaller *a* is expected for the newer data-set as only mammography-diagnosed tumours are included in the newer data [11]. A limitation of the tumour volume model is that we considered in this work only breast cancer data. However, the intent of this work was a proof of principle and detailed analysis of tumour volume distributions for various tumour sites is in preparation.

If exponential tumour growth is used as a transformation function the probability density function for tumour volumes can be transformed into the probability density function of tumour growth or lag times, respectively. The obtained lag time distribution function can be used in the development of mechanistic models of background cancer risk. Usually, such models combine the probability of occurrence of a malignant cell per individual per year with a constant lag time in order to estimate cancer induction [12–16]. Replacing a constant lag time by a lag time distribution may improve the quality of such models. We would like to note that the average lag time for breast cancer shown in Figure 3 depends strongly on the tumour growth factor *b* which we have taken from literature. The goal of this work was not to determine the absolute lag times for breast cancer, but to derive an expression for the distribution of lag times.

A limitation of this work is that tumour growth was simply modeled with an exponential model. An exponential model might be a satisfying approximation as long as the tumours are small. However, it is known that the exponential model is not universally appropriate and many other models including the Gompertz model have been proposed [19]. It might be interesting to apply the formalism of this work to other tumour growth models and to study its impact on the tumour size distribution.

It was shown that the assumed exponential variation of tumour volumes can be used to derive an analytical representation for the population averaged TCP. The Poisson based TCP model for individuals was integrated by using the volume distribution as a weighting function. This resulted in the logistic function as a population averaged TCP model. The logistic function was used in the past to fit successfully observed tumour control data for radiotherapy patients without any bio-physical justification. In this work a logistic TCP model for cohorts of patients is explicitly derived from a Poisson TCP-model for individuals.

The methodology presented in this work enables in principle the cohort TCP model given by equation 9 to be fitted to survival data for groups of patients. However, radiotherapy patients who have been similarly treated include not only different tumour volumes, but also different clonogen densities, different intrinsic radiosensitivites and different oxygenation, none of which were considered in this work. We are currently working on an integration of most of these factors into our analytical model. Therefore, this work should be viewed as a first step towards the development of a TCP model that can be applied equally to individuals and to populations of patients.

## Data Availability

All data referred to in the manuscript are available from the author.

## Footnotes

† By tumour size we mean here the size of the tumour when the treatment starts.

## Notes

### Competing Interest Statement

The authors have declared no competing interest.

### Funding Statement

No external funding was received.

### Author Declarations

No ethics committee approvals have been obtained.

